# Exploring the creation or adaptation of knowledge mobilization products for culturally and linguistically diverse audiences: a scoping review

**DOI:** 10.1101/2023.09.06.23295083

**Authors:** Sarah A Elliott, Shannon D Scott, Liza Bialy, Kelsey Wright, Lisa Hartling

**Author notes:** **Contact Author** Dr. Sarah Elliott 4-474 Edmonton Clinic Health Academy 1405 87th Ave Edmonton, AB Canada T6G 1C9.

## Abstract

****Introduction**:** Connecting end-users to research evidence has the power to improve patient knowledge and inform health decision-making. However, recognized barriers to or determinants of effective knowledge mobilization (KMb) are differences in culture and language among the end users of the evidence. This scoping review set out to understand current processes and practices when creating or adapting KMb products for culturally and linguistically diverse (CALD) audiences.

****Methods**:** We searched 3 databases (Ovid Medline, CINAHL via EBSCOhost, PsychINFO) from 2011 until August 2021. We included any literature about KMb product creation or adaptation processes serving CALD communities. A primary reviewer screened all identified publications and a second reviewer screened publications excluded by the primary. Data were extracted using a standardized form by one reviewer and 10% were verified by a second reviewer. Studies were categorized by type of adaptation (‘surface’ or ‘deep’ structure) and mapped based on type of stakeholder engagement used (i2S model). A search update was run in July 2023, and screening and extraction are in progress.

****Results**:** Seven thousand four hundred and five unique titles and abstracts were reviewed, 319 full-text studies were retrieved and reviewed, and 24 studies were included in final data extraction and mapping. Fifteen studies (63%) created or adapted exclusively text-based KMb products such as leaflets and pamphlets and 9 (38%) produced digital products such as videos (n=4, 16%), mobile applications (n=3, 13%), website (n=1, 4%) and a CD ROM (n=1, 4%). Eight studies (33%) reported following a framework or theory for their creation or adaptation efforts. Only five studies (21%) demonstrated ‘deep structure’ cultural sensitivity and applied all five (Inform, Consult, Involve, Collaborate, and Support) levels of stakeholder engagement. Four (17%) studies included reflections from the research teams on the processes for creating or adapting KMb products for CALD communities.

****Conclusion**:** Included studies cited a variety of methods in creating or adapting KMb products for CALD communities. Successful uptake of created or adapted KMb products was often the result of collaboration with end-users for more applicable, accessible and meaningful products. Further research developing guidance and best practices is needed to support the creation or adaptation of KMb products with CALD communities.

## Introduction

Knowledge mobilization (KMb) (an umbrella term encapsulating knowledge translation, knowledge transfer, and knowledge exchange) ^1^ is based on communicating scientific evidence to relevant end-users^2^. Within healthcare, KMb products (e.g. patient decision aids, educational resources) can help patients and families in their healthcare decision-making to improve health outcomes and reduce health system costs. ^3^ Successful uptake of evidence is contingent on relevance of the KMb products for the target end-user. ^2^ However, some recognized barriers or determinants of effective KMb are differences in culture and language among the end-users of the evidence.

While KMb efforts have advanced substantially in the field of health promotion over the last decade, predominant cultures often comprise the accessible pool of engaged end-users. ^4, 5^ Subsequently, KMb products frequently entail English communication, and mainstream images not conveying relevance to minority cultures. Similarly, most KMb products assume end-users possess a certain level of health literacy and are relatively familiar with their healthcare system, which may not represent the experiences of many newcomer cultural groups. Public health agencies (e.g. Health Canada) sometimes provide linguistic translations of healthcare information for common languages; however, linguistic translation does not guarantee accessibility and relevance of healthcare information for the target communities. Instead, nuanced visuals, relevant terms, and overall cultural sensitivity have proved more desirable for end-users. ^6^

Resnicow and colleagues^7^ have proposed that cultural adaptation consists of two dimensions: surface structure and deep structure. Aspects of culture that are easily observable to external onlookers like language, clothing, and ethnicity would fall under surface structure, while historical and psychological influences on health decisions would fall under deep structure. Though admittedly gray in nature, the delineation of surface and deep aspects provides potential categories of cultural adaptation. In terms of the application of cultural sensitivity, efforts to develop or adapt KMb products could include surface structures of language and appearance of end-users, as well as deep structures of historical barriers and psychological stressors for end-users. ^8^ Resnicow and colleagues suggest that both surface and deep structures of cultural knowledge are essential for well-rounded cultural adaptations, and encourage the involvement of end-users to understand the nuanced aspects.

Types and extent of end-user engagement can also vary in KMb product creation and adaptation.^9–11^ Bammer ^12^ proposed a modified version of the International Association for Public Participation (IAP2) stakeholder engagement model, where researchers are positioned as support for the directions chosen by end-users, rather than decision-makers themselves. The positioning of end-users as experts in their own information needs and preferences mirrors other public participation frameworks often employed by heath researchers (e.g. Participatory Action Research (PAR), ^13^ Community-Based Participatory Research (CBPR), ^14^ etc.). Both cultural adaptation and end-user engagement appear to be core pillars in KMb product creation and adaptation.

While there are several processes (e.g. translation and cultural brokerage, ecological validity model) of cultural adaptation that have been previously applied to adapt health intervention programs, ^15^ and patient reported outcome scales, ^16^ no guidance currently exists on how best to develop or adapt KMb products that reach diverse end-user needs. As an initial step towards creating a unified, effective KMb product creation or adaptation best practices, this scoping review (ScR) aimed to map what approaches researchers have used to culturally and linguistically adapt KMb products. The following questions guided this scoping review:

## Key questions

1. What *approaches* have researchers used when creating or culturally adapting KMb products for culturally and linguistically diverse (CALD) end-users?
2. What are the *key consideratio*ns when creating or culturally adapting KMb products for CALD end-users?

Understanding what methods have previously been used, resources required, as well as key considerations for how best to create or adapt KMb products will support researchers and healthcare organizations in developing or adapting effective resources for CALD communities.

## Methods

### Review methods

This ScR followed the methodological framework proposed by Arksey and O’Malley, ^17^ enhanced by Levac et al. ^18^ Specifically, we followed these six steps: (1) identifying the research question; (2) identifying the relevant studies; (3) study selection; (4) charting the data; (5) reporting the results and (6) consultation. Reporting of the review adheres to the Preferred Reporting Items for Systematic Reviews and Meta-Analyses (PRISMA) – 2018 Extension for Scoping Review. ^19^

### Search strategy

In collaboration with a research librarian and content experts, we developed and refined a comprehensive search strategy. The strategy combined subject headings and keywords for terms related to KMb, knowledge exchange, knowledge mobilization, cross-culture, culturally appropriate, CALD communities, and adaptation of health information and implementation science. On August 12, 2021 we searched Ovid Medline (1946-), CINAHL via EBSCOhost (1937-), PsycINFO (2002-), as well as ProQuest Dissertations & Theses Global to identify grey literature. Search results were exported to EndNote V.X7 (Clarivate Analytics) and duplicates removed before the file was provided to reviewers for screening in Microsoft Excel. The search was limited to English language, peer-reviewed studies published in academic journals from 2011 to August 2021. A search update was run in July 2023, screening and data extraction for the update is in progress.

### Study selection

All published study design types and secondary evidence syntheses were included if they contained a patient, public or consumer population and culturally created or adapted a KMb product. We defined a KMb product as any health research-based product that supported decision-making to provide explicit recommendations, and/or meet knowledge needs. We excluded any studies that only performed purely linguistic translations of a KMb product or the validation of translated measurement tools/questionnaires. Interventions without standalone KMb products for end-user decision making were also excluded.

### Screening

One reviewer screened titles and abstracts of each study as “include/unsure” or “exclude” based on a priori inclusion criteria (Supplementary Material 1). A second independent reviewer verified all studies excluded by the first reviewer. Both reviewers performed a pilot screen where they independently screened 10% of the studies to assess consistency. Two independent reviewers reviewed the full-text of each included study from the primary screening. When agreement on a citation or full-text could not be reached between two reviewers, a third senior reviewer was consulted for resolution.

### Data extraction

The following details were extracted from each study: publication characteristics, study design, population, KMb product description, methods of creation or adaptation, stakeholder engagement processes, KMb product evaluation processes, and reflections from researchers. Data were collected using a standardized form by one reviewer and verified by a second reviewer. Any discrepancies were resolved through a third-party decision.

### Data analysis

We performed a narrative synthesis, guided by a qualitative content analysis approach^20^ to summarize the quantity, content, and coverage of the evidence including summary statistics on studies examining the different ways of culturally developing or adapting KMb products. Processes for creation or adaptation were mapped into five categories (Product creation, Literature search, Stakeholder engagement, Resources utilized, Evaluation) representing different methods reported throughout the literature.

Cultural adaptations were categorized into two broad groups, surface and deep structure. ^7, 21^ Surface structure involved coordinating materials and messages to observable characteristics of the target population (i.e., imagery, sounds, backgrounds, clothing, etc.). Deep structure involved contextualizing the social, historical, environmental, and psychological features of the proposed end-user group. Additionally, Bammer’s iS2 version^12^ of the IAP2 Spectrum of Public Participation was used to classify levels of engagement across each study. ^22, 23^ The i2S is a spectrum of engagement across five stages: informing stakeholders of health information and research processes, consulting with stakeholders for relevant cultural considerations, involving stakeholders throughout the research processes, collaborating with stakeholders in research decisions, and supporting stakeholders in designing and implementing appropriate and desirable research and dissemination methods.

## Results

### Search results

The search strategy (Supplementary Material 2) captured 9,677 studies. After removing duplicates, 7,405 unique studies were screened by title and abstract. Of these, the full-text of 319 were reviewed and 24 met eligibility criteria and were included in the review. The PRISMA flow diagram (Figure 1) provides a detailed outline of the screening and selection process.

**Figure 1:**
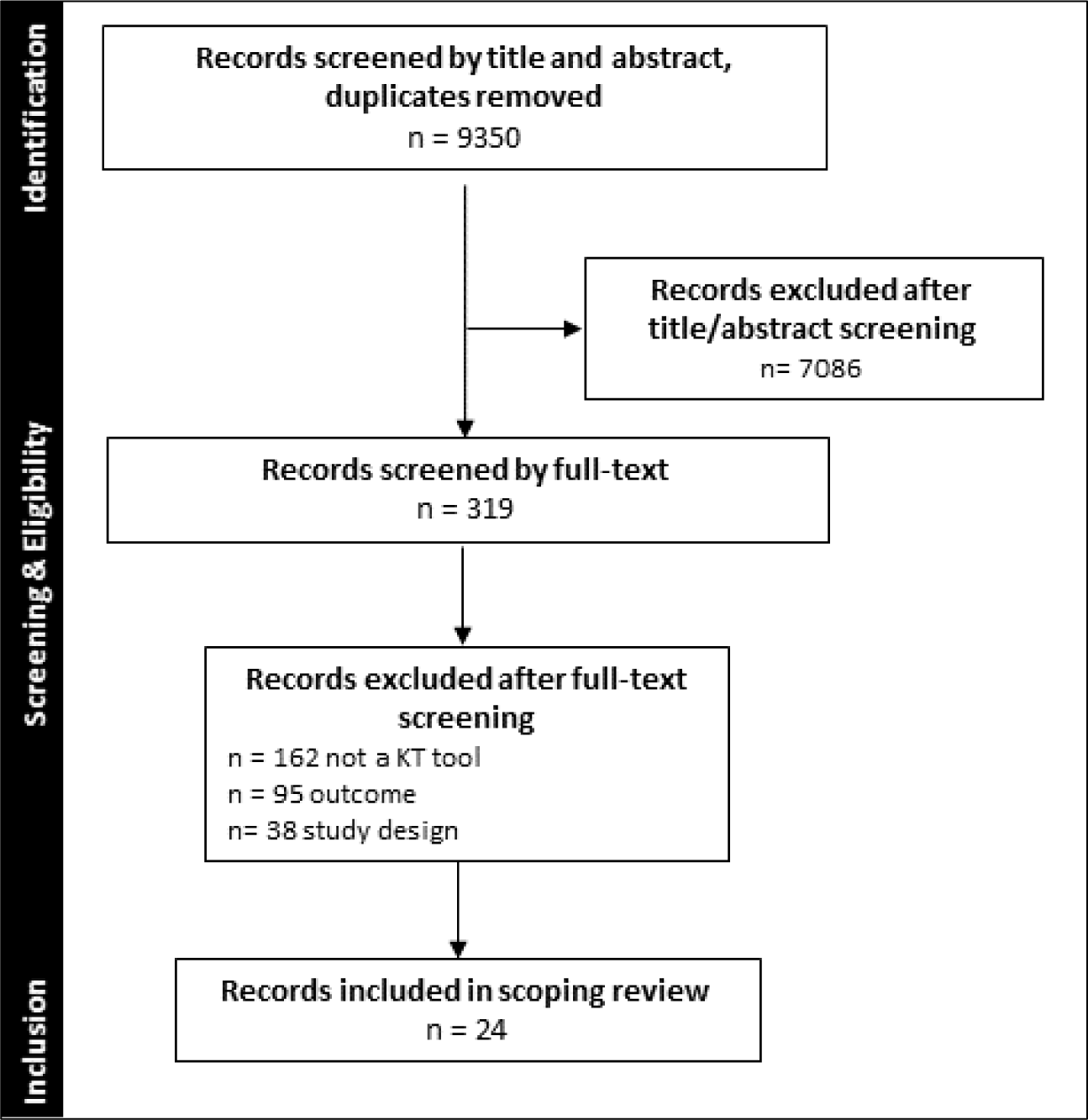
PRISMA flow diagram.

### Study characteristics

Table 1 provides a summary of the included studies’ characteristics. The majority of studies were from USA (54%, n=13) and Canada (13%, n=3) with a single study from each of the following countries: United Kingdom, Portugal, Norway, Israel, United Arab Emirates, Denmark, New Zealand, and Egypt. Most of the studies used qualitative methods (83%, n=20) followed by mixed methods (13%, n=3) and quantitative methods (8%, n=2). Fourteen (58%) reported the age of participants, most (n=12, 86%) focused on adult populations, two (14%) on adolescents, and 10 (42%) did not report on age.

**Table 1:** Characteristics of studies culturally creating or adapting KMb products.

| Study Characteristics |  |  | Population Characteristics |  | KT Product Characteristics |  |
| --- | --- | --- | --- | --- | --- | --- |
| Author<br>Year<br>Country | Study design /<br>methods<br>Sample size | Objective and setting | Age,<br>mean±SD<br>(range)*<br>Males, n<br>(%) | Primary<br>language<br>Race<br>Condition | KT product<br>description and format | Target<br>end-users |
| Abbas-Dick<br>2018<br>Canada | • Qualitative<br>• Participatory research with content analysis<br>• N=11 | To work in partnership with Indigenous communities to create an eHealth breastfeeding resource for Indigenous families | 31 (18-57)<br>0 (0) | • English<br>• Indigenous<br>• Breastfeeding | Online breastfeeding co-parenting resources including text, video, games, quizzes, and links to internet resources. | Public<br>(families) |
| Ali<br>2018<br>UK | • Qualitative<br>• Interviews<br>• N=20 | To develop a simple health literacy intervention aimed at supporting informed reproductive choice among members of UK communities practicing customary consanguineous marriage | NR<br>NR | • Urdu<br>• South Asian<br>• Consanguineous marriage | Simple health literacy leaflet on reproductive choices. | Public |
| Arnold<br>2011<br>USA | • Qualitative<br>• Focus groups<br>• N=13 | To describe how the desire of an American Indian community in Wyoming grew into an American Indian/Alaska Native women's advisory committee, a culturally appropriate prenatal education booklet, and | NR<br>NR | • Various<br>• Indigenous<br>• Prenatal education | A comprehensive prenatal booklet that would "tell the story of the way to have a healthy pregnancy." Presented in a conversational format. | Parents |
| Author<br>Year<br>Country | Study design /<br>methods<br>Sample size | Objective and setting | Age,<br>mean±SD<br>(range)*<br>Males, n<br>(%) | Primary<br>language<br>Race<br>Condition | KT product<br>description and format | Target<br>end-users |
|  |  | a national initiative for a non-profit organization |  |  |  |  |
| Baptista<br>2020<br>Portugal | • Qualitative<br>• Interviews<br>• N=15 | To translate and culturally adapt an English language patient decision aid addressing prostate cancer screening, so it can be used by Portuguese men. | 61±4.9 (55-69)<br>15 (100) | • Portuguese<br>• White<br>• Prostate cancer | Prostate cancer screening text-based decision aid. | Patients |
| Best<br>2012<br>USA | • Mixed methods<br>• Focus groups, interviews, RCT<br>• N=200 | To work with a group of African American women to 1) identify important spiritual elements to be included in health communication materials, and develop a spiritually-framed Breast Cancer Screening (BCS) message in response to their feedback and 2) evaluate the effectiveness of the spiritually-framed BCS | 48.9±15.4 (25-88)<br>0 (0) | • English<br>• African American<br>• Breast cancer screening | Print and radio-based messaging for breast cancer screening information. | Patients |
| Author<br>Year<br>Country | Study design /<br>methods<br>Sample size | Objective and setting | Age,<br>mean±SD<br>(range)*<br>Males, n<br>(%) | Primary<br>language<br>Race<br>Condition | KT product<br>description and format | Target<br>end-users |
|  |  | message compared to a more traditional BCS message among African American women using a cognitive response analysis |  |  |  |  |
| Bilbrey<br>2018<br>USA | • Mixed methods<br>• Focus groups, online surveys<br>• NR | To develop materials which help answer questions and support the decision-making process for families related to brain donation | NR<br>NR | • Spanish<br>• Latino<br>• Brain donation | Bilingual brochure highlighting the benefits and process of brain donation. | Patients (families) |
| Drago<br>2018<br>USA | • Qualitative<br>• Interviews<br>• N=22 | To characterize Latino parental perceptions of antenatal counseling in order to construct and validate a Spanish decision-aid to improve parental knowledge of prematurity after antenatal consults | NR<br>3 (14) | • Spanish<br>• Latino<br>• Premature birth | Card-based English decision-aid for parents experiencing premature birth. | Patients |
| Gordon<br>2015<br>USA | • Qualitative<br>• Focus groups<br>• N=76 | To describe the community engagement | NR<br>NR | • Spanish<br>• Latino | Website on routine transplant education. | Patients |
| Author<br>Year<br>Country | Study design /<br>methods<br>Sample size | Objective and setting | Age,<br>mean±SD<br>(range)*<br>Males, n<br>(%) | Primary<br>language<br>Race<br>Condition | KT product<br>description and format | Target<br>end-users |
|  |  | processes and data collection efforts used to identify culturally appropriate content and describe development of website content and design, and usability testing. |  | • Kidney donation and transplantation |  |  |
| Grasaas<br>2019<br>Norway | • Qualitative<br>• Interviews<br>• N=11 | To describe the translation and cultural adaptation of the iCanCope with Pain mobile app into the Norwegian context and evaluate the app's usability using a phased approach at the University of Agder laboratory | NR (16-18)<br>NR | • Norwegian<br>• White<br>• Adolescent pain | Mobile app containing resources for current best practices for pain self-management. | Patients |
| Grinker<br>2015<br>USA | • Qualitative<br>• Interviews<br>• N=23 | To describe a methodology for translating outreach materials for Autism Spectrum Disorder using the Autism Speaks First 100 Days Kit as an exemplar. | NR<br>NR | • Korean<br>• East Asian<br>• Autism spectrum disorder | Autism Speaks First 100 Days Kit: text document. | Patients<br>(families) |
| Author<br>Year<br>Country | Study design /<br>methods<br>Sample size | Objective and setting | Age,<br>mean±SD<br>(range)*<br>Males, n<br>(%) | Primary<br>language<br>Race<br>Condition | KT product<br>description and format | Target<br>end-users |
| Guttman<br>2013<br>Israel | • Qualitative<br>• Interviews<br>• N=104 | To develop and present health rights information materials for a disadvantaged cultural minority, the Ethiopian immigrant community in Israel. | NR (NR)<br>44 (42) | • English<br>• Middle Eastern/North African<br>• Health rights information | Two styles of video: 1 video with a humorous storyline format, and 2 videos with a narrator relaying serious information. Two styles of print materials: a photonovella, and an illustrated booklet. | Public |
| Harvey<br>2011<br>USA | • Qualitative<br>• Interviews<br>• N=20 | To develop, produce and disseminate culturally and linguistically appropriate health brochures in community health center serving populations of low health literacy, in Philadelphia | NR (20-60)<br>NR | • Spanish<br>• Latinx<br>• Health information topics | Written health brochures on common health information to promote health literacy. | Public |
| Hashim<br>2013<br>UAE | • Mixed methods<br>• Focus groups, survey<br>• N=17 | To investigate the content and design preferences of printed health education leaflets among Arabic communities, conducted in clinic | 47.4±14.3<br>(17-70)<br>9 (53) | • Arabic<br>• Middle Eastern/North African<br>• Preferences for health education materials | Diabetes health education materials in a trifold brochure. | Public |
| Author<br>Year<br>Country | Study design /<br>methods<br>Sample size | Objective and setting | Age,<br>mean±SD<br>(range)*<br>Males, n<br>(%) | Primary<br>language<br>Race<br>Condition | KT product<br>description and format | Target<br>end-users |
|  |  | waiting areas at public hospital |  |  |  |  |
| Hempler<br>2015<br>Denmark | • Qualitative<br>• Design-based research approach<br>• N=18 | To develop culturally sensitive dialog tools to support person-centered dietary education targeting Pakistani immigrants in Denmark with type 2 diabetes | NR<br>NR | • English<br>• South Asian<br>• Type 2 diabetes | Diabetes dialog tools including interactive engagement, online videos, and text. | Patients |
| Lee<br>2019<br>USA | • Qualitative<br>• Focus groups<br>• N=20 | To demonstrate how a culturally targeted and tailored mobile text messaging intervention, mobile screening (mScreening), was developed to promote the uptake of Papanicolaou tests and human papillomavirus vaccine (HPV) among young Korean American immigrant women | 26 (21-29)<br>0 (0) | • Korean<br>• East Asian<br>• Cervical cancer | HPV mScreening mobile app. | Public |
| Li<br>2012<br>USA | • Qualitative<br>• Interviews<br>• N=5 | To develop, pilot test and evaluate an innovative, culturally | 74.3±4.8<br>(71-81)<br>4 (20) | • Mandarin<br>• East Asian<br>• Hypertension | Hypertension CD-ROM including visual aids and audio features. | Patients |
| Author<br>Year<br>Country | Study design /<br>methods<br>Sample size | Objective and setting | Age,<br>mean±SD<br>(range)*<br>Males, n<br>(%) | Primary<br>language<br>Race<br>Condition | KT product<br>description and format | Target<br>end-users |
|  |  | relevant CD-ROM with a focus on hypertension education and management, directed at the older Chinese immigrant population, at Asian health clinics in San Francisco |  |  |  |  |
| Mathieson<br>2012<br>New Zealand | • Qualitative<br>• Interviews<br>• N=16 | To adapt an existing cognitive behavioural therapy-based, guided self-management intervention for near-threshold mental health syndromes in primary care, for Maori, and to examine its acceptability and effectiveness. | 38.9 (20-65)<br>9 (19) | • te reo (Maori language)<br>• Indigenous<br>• Mental health syndromes | Self-management booklets on relevant mental health issues. | Patients |
| Meherali<br>2021<br>Canada | • Mixed methods<br>• Focus groups, surveys<br>• N=68 | To (1) develop Acute Otitis Media KT tools for Canadian parents, (2) adapt the KT tools across cultural contexts, and (3) evaluate the usability of the adapted KT tools. | NR (0-50)<br>4 (6) | • Urdu<br>• South Asian<br>• Acute otitis media | Digital arts-based KT tools on how to manage acute otitis media. | Parents |
| Author<br>Year<br>Country | Study design /<br>methods<br>Sample size | Objective and setting | Age,<br>mean±SD<br>(range)*<br>Males, n<br>(%) | Primary<br>language<br>Race<br>Condition | KT product<br>description and format | Target<br>end-users |
| Norris<br>2021<br>USA | • Qualitative<br>• Focus groups<br>• N=25 | To create a multicultural adaptation of the Mighty Girls program using a mobile app that is less costly to disseminate and is acceptable to parents of 7<sup>th</sup> grade girls | NR (10-14)<br>0 (0) | • English<br>• Latinx<br>• Sexual health | A narrative-generating app to support sexual health behavior change. | Patients<br>(families) |
| Rami<br>2018<br>Egypt | • Quantitative<br>• RCT<br>• N=20 | To translate and culturally adapt the Behavioral Family Psycho-Education Program and conduct a preliminary efficacy evaluation for outpatients suffering from schizophrenia | NR (18-65)<br>NR | • Arabic<br>• Middle Eastern/North African | Informational leaflets about schizophrenia, high emotionally expressive families, and training materials for communication and problem-solving skills. | Patients<br>(families) |
| Sharpe<br>2013<br>USA | • Qualitative<br>• Interviews<br>• N=32 | To describe the process and application of formative assessment with women attending an American Indian women's clinic to the development and pretesting a culturally tailored brochure. | 41 (20-61)<br>0 (0) | • English<br>• Indigenous<br>• Women with human papillomavirus | Informational brochure with the HPV vaccine recommendations, its availability, and accessibility. | Patients |
| Author<br>Year<br>Country | Study design /<br>methods<br>Sample size | Objective and setting | Age,<br>mean±SD<br>(range)*<br>Males, n<br>(%) | Primary<br>language<br>Race<br>Condition | KT product<br>description and format | Target<br>end-users |
| Stanley<br>2018<br>USA | • Qualitative<br>• Focus groups<br>• N=24 | To describe formative research undertaken to guide adaptation for AI youth of a prevention intervention, Be Under Your Own Influence (BUYOI), previously found to be effective in reducing substance use among middle-school youth. | NR<br>NR | • English<br>• Indigenous<br>• Substance use | BUYOI paper-based messages, targeted to middle-school youth. | Public |
| van der Steen<br>2013<br>Canada | • Qualitative<br>• Content analysis of transcribed materials<br>• NR | To develop a decision-making booklet on palliative care issues. To understand which aspects are sensitive from an ethical and cultural point of view within the respective cultural contexts in giving shape to palliative care in dementia | NR<br>NR | • Dutch, Japanese and Italian<br>• White/East Asian<br>• Dementia | A booklet on comfort care at the end of life with dementia. | Patients (families) |
| Van Son<br>2014<br>USA | • Qualitative<br>• Interviews<br>• N=10 | To describe a project in which culturally targeted diabetes education materials for | NR (over 65)<br>NR | • Russian<br>• White<br>• Diabetes | 12 educational documents created with Russian on one side and English on the other to | Patients |
| Author<br>Year<br>Country | Study design /<br>methods<br>Sample size | Objective and setting | Age,<br>mean±SD<br>(range)*<br>Males, n<br>(%) | Primary<br>language<br>Race<br>Condition | KT product<br>description and format | Target<br>end-users |
|  |  | older Russian-speaking<br>immigrants were<br>designed and<br>developed, delivered at<br>diabetes clinics |  |  | support patients make<br>better health decision<br>with regards to food. |  |

\* age in years. KMb, knowledge translation; UK, United Kingdom; RCT, randomized controlled trial

End-user groups varied across the included studies, with Indigenous English speakers (17%, n=4) and Latinx Spanish speakers (17%, n=4) as the most frequently reported cultural communities served. Health topics of the KMb products included: diabetes (13%, n=3), organ donation (8%, n=2), mental health conditions (8%, n=2), prenatal care (8%, n=2), cancer (8%, n=2), HPV vaccine (8%, n=2), health literacy (8%, n=2), and acute otitis media, Autism Spectrum Disorder, breastfeeding, dementia, hypertension, pain,, reproductive choice, sexual health, substance use each presented in a single study. Fifteen studies (63%) created or adapted exclusively non-digital KMb products such as leaflets and pamphlets and 9 (38%) produced digital products such as videos (n=4, 16%), mobile applications (n=3, 13%), website (n=1, 4%) and a CD ROM (n=1, 4%).

### Knowledge Product Creation

Full details of reported creation or adaptation processes are outlined in Table 2.

**Table 2:**
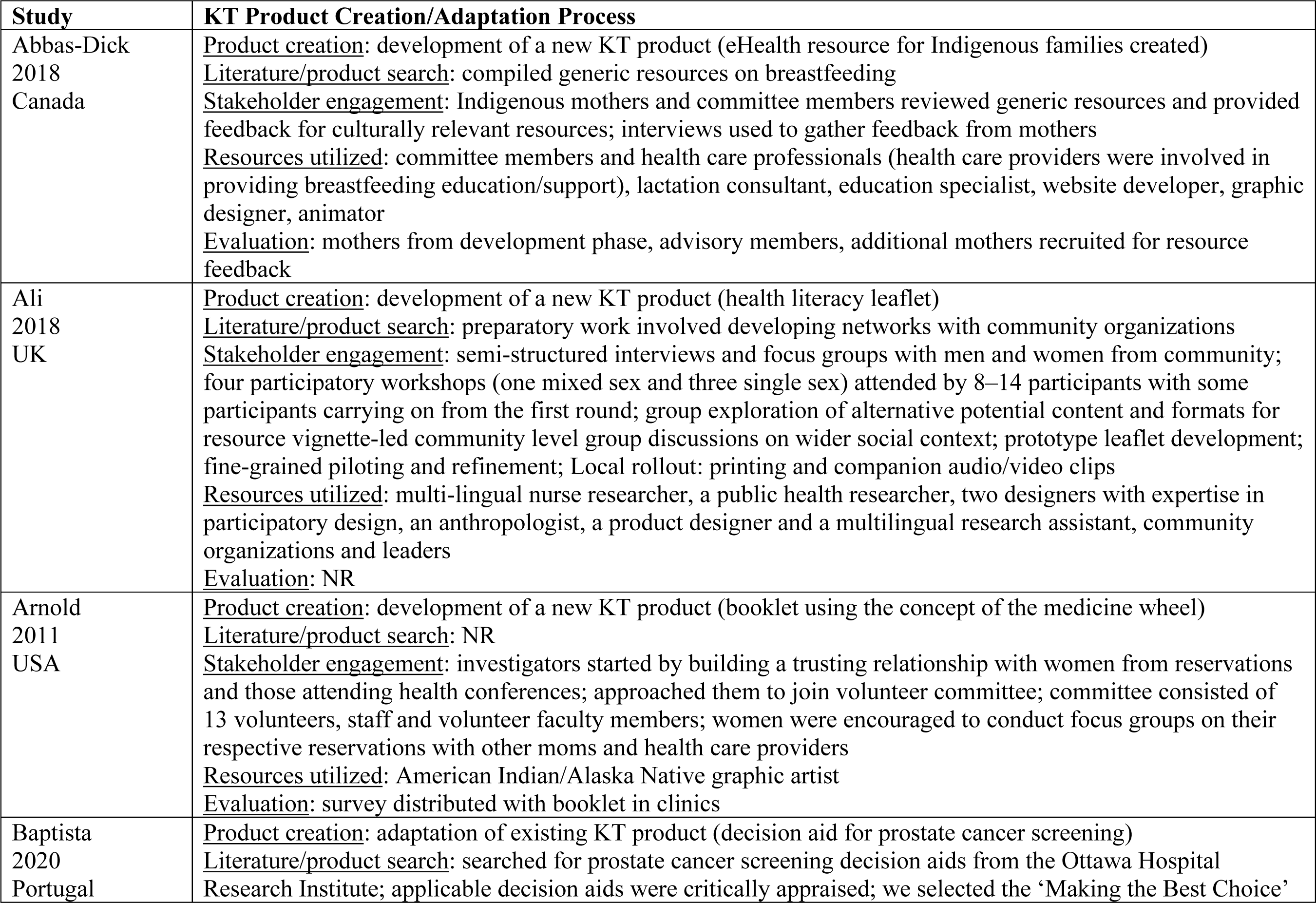
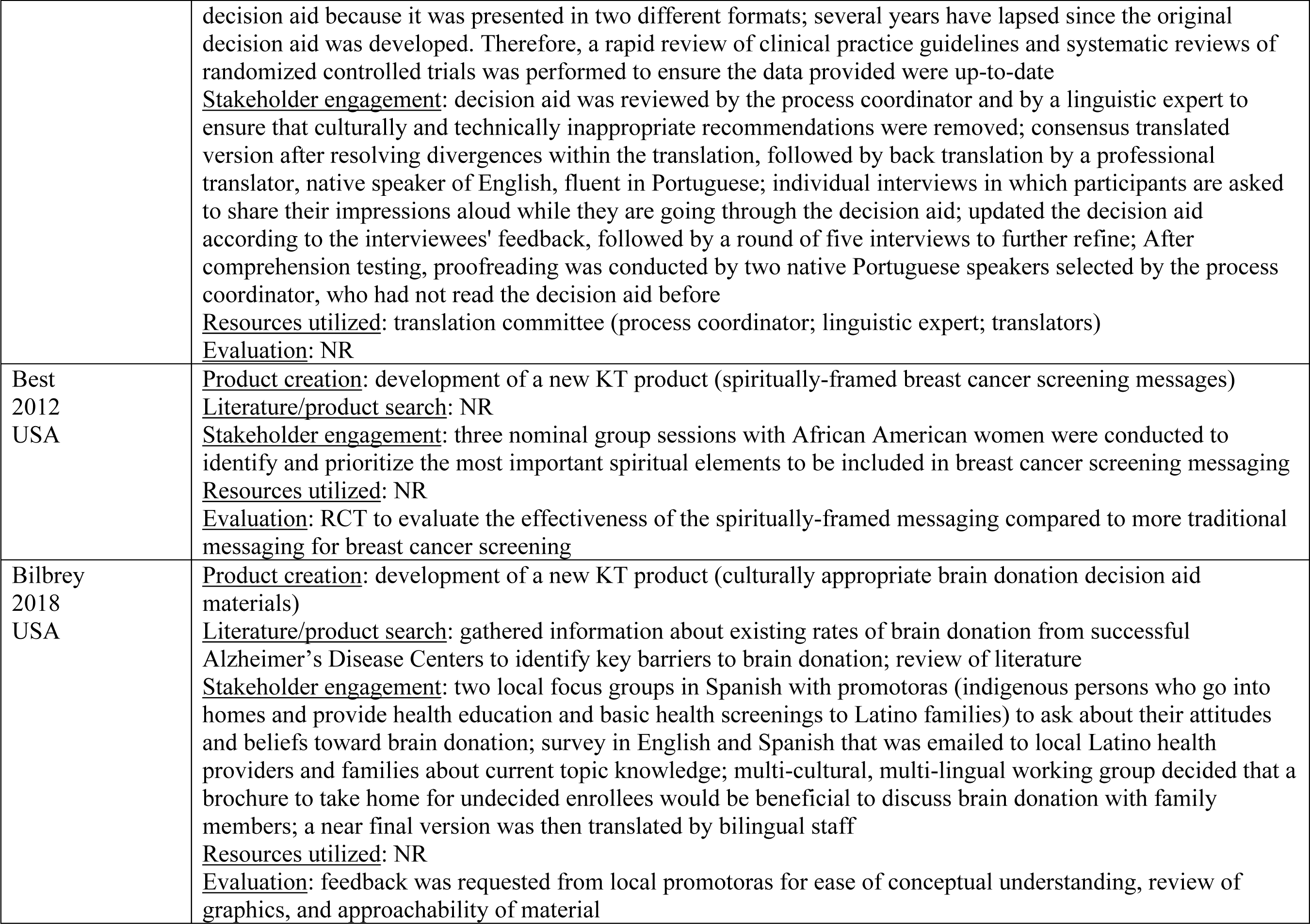
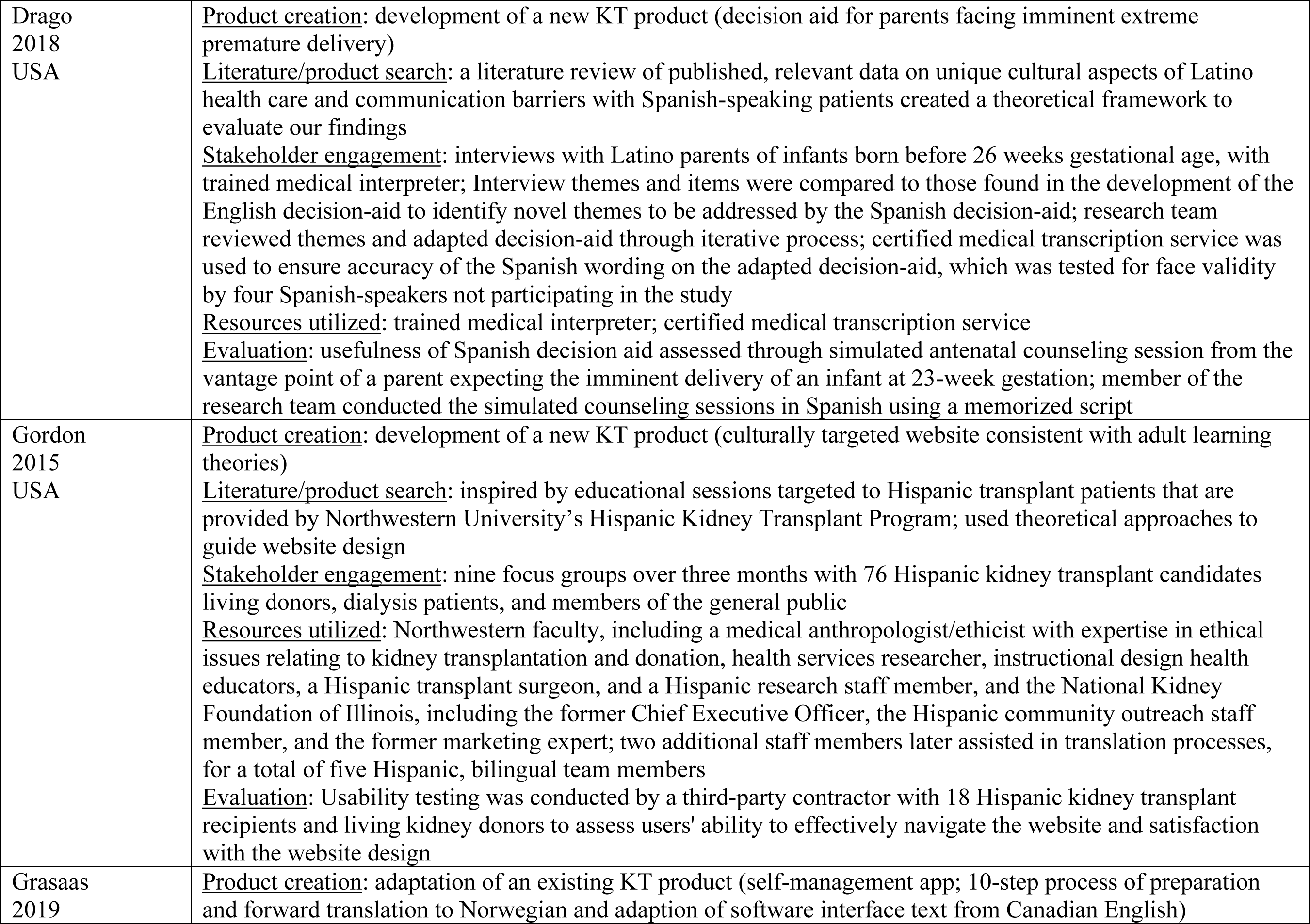
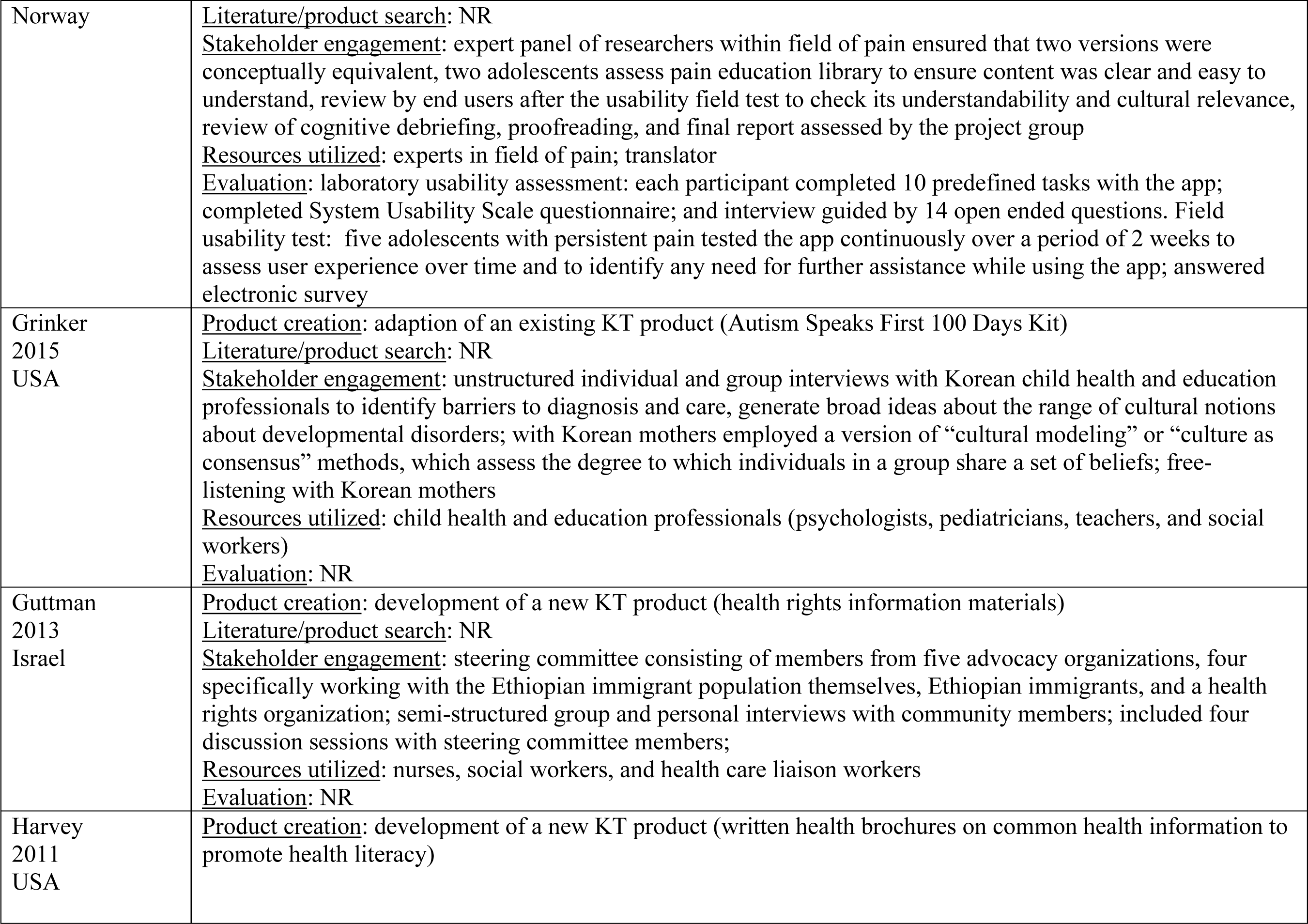
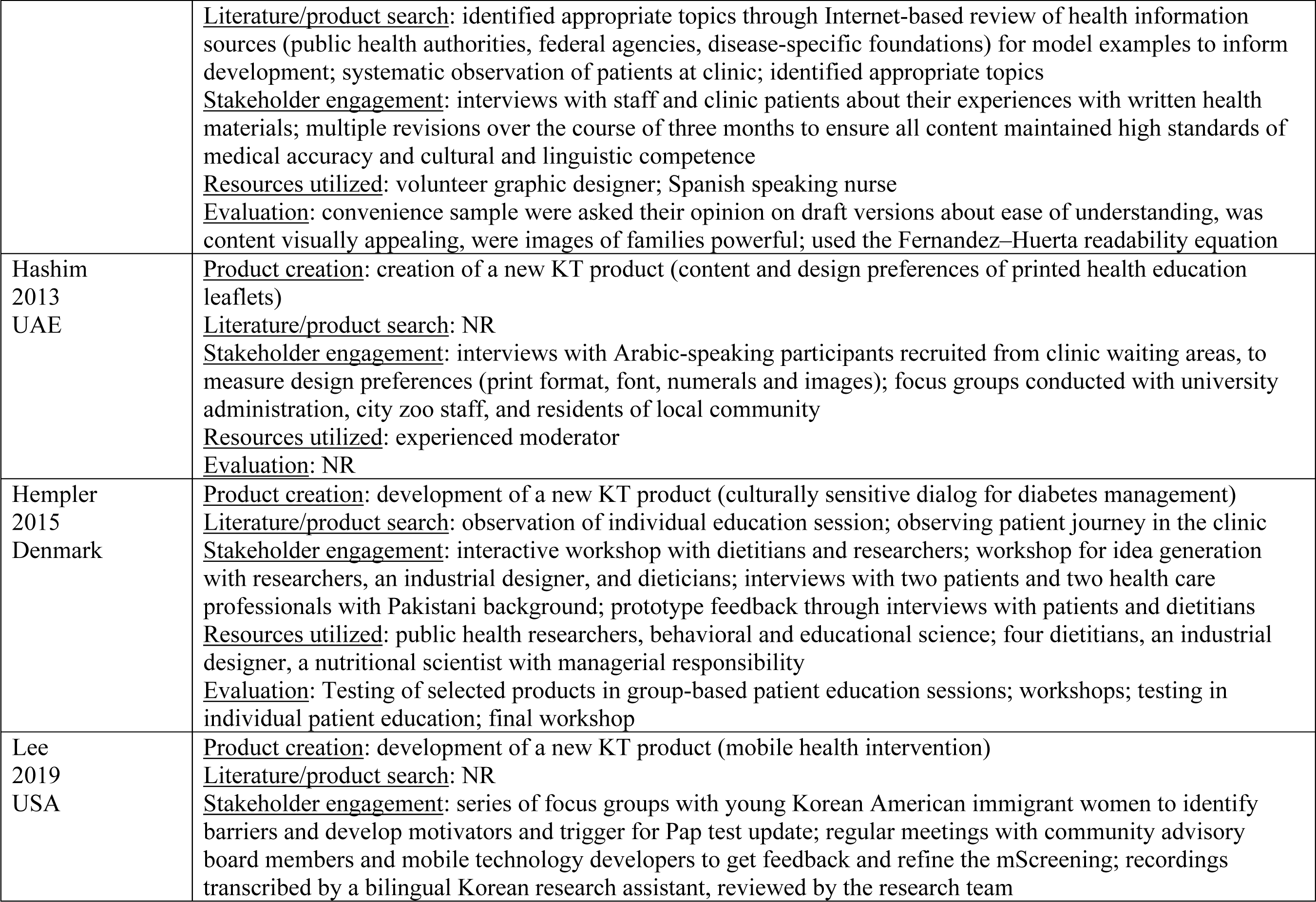
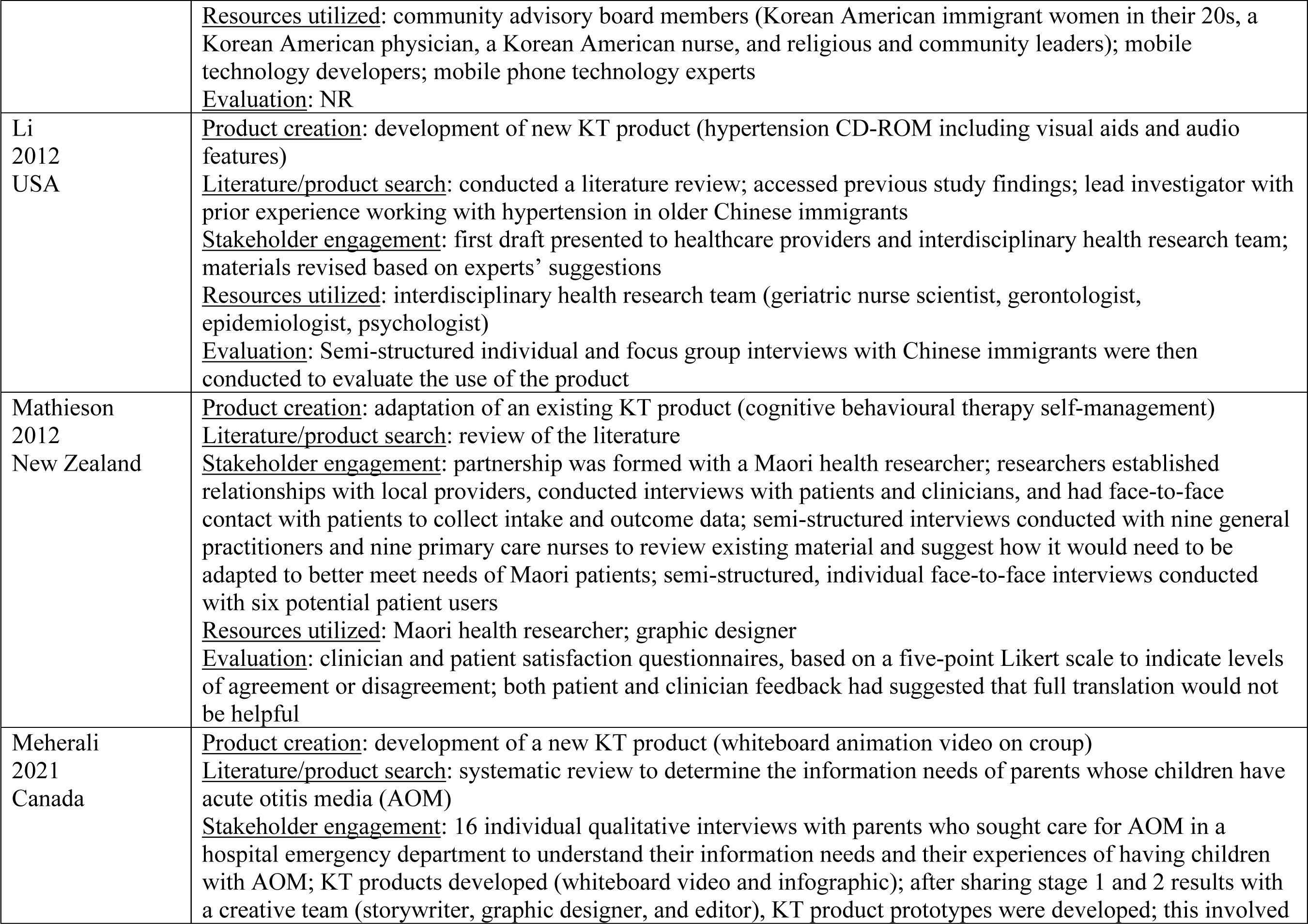
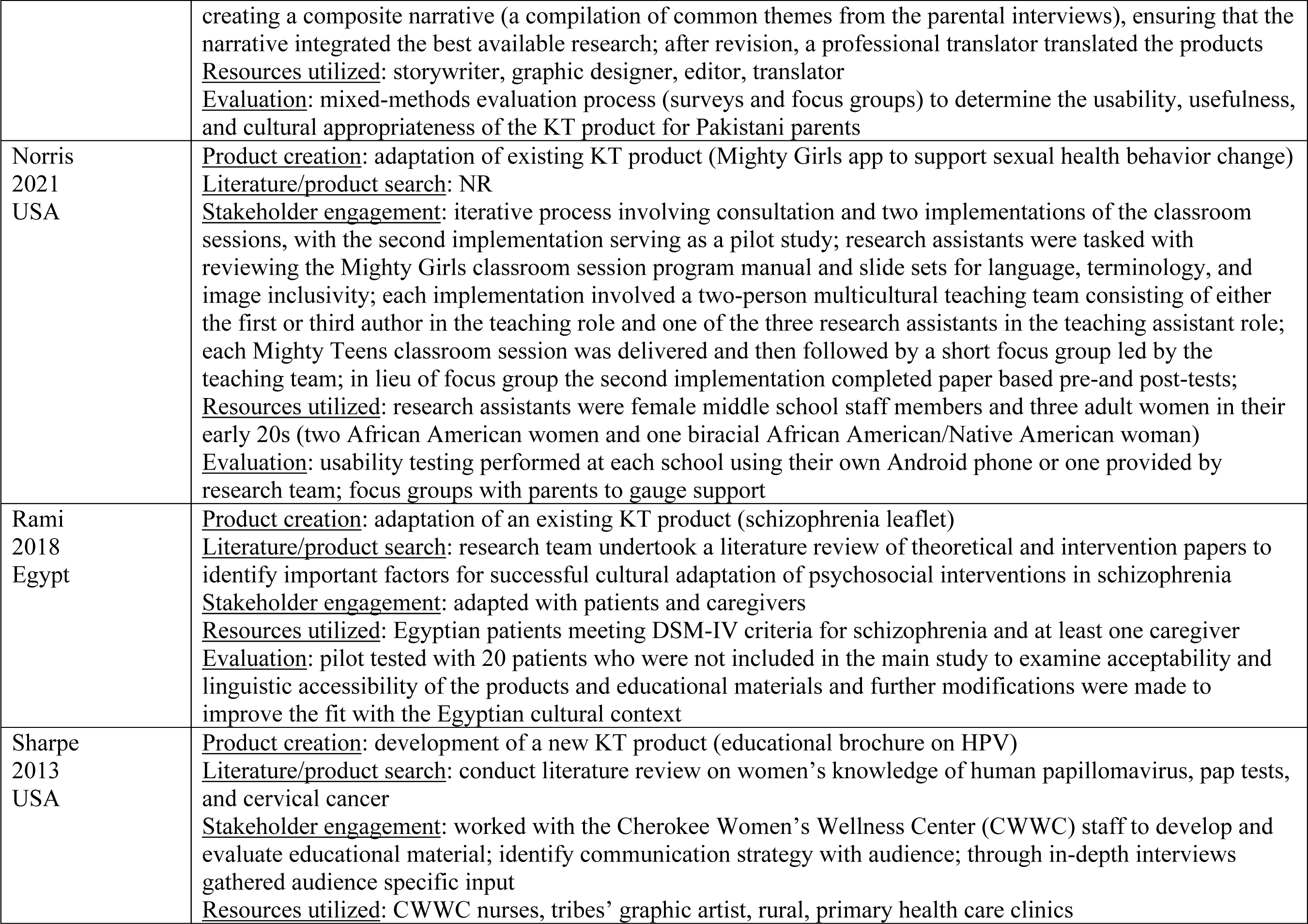
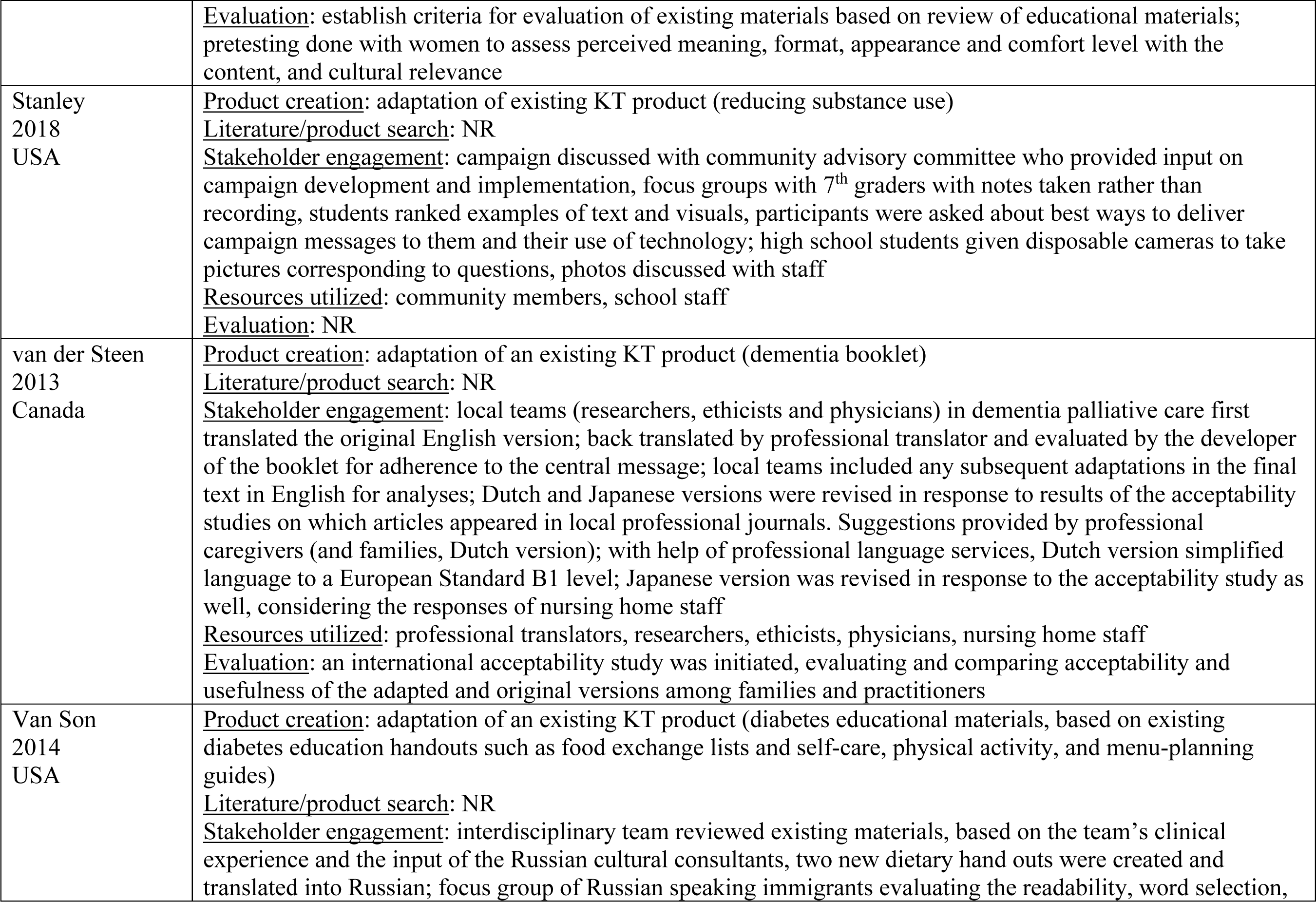
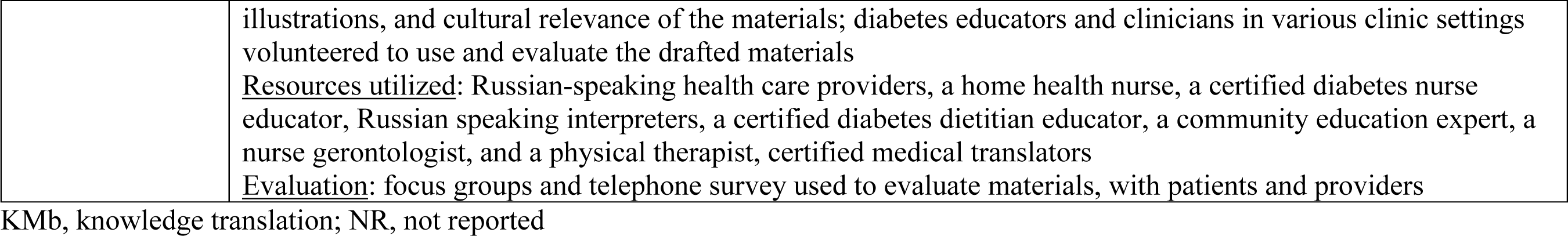
Processes of creating or adapting of KMb products.

#### Creation or Adaptation Processes

Each included study followed unique processes for developing or adapting a KMb product for CALD communities. Thirteen studies (54%) reported the processes of developing a new KMb product, while eleven studies (46%) outlined their process for adapting a pre-existing KMb product for a specific end-user group. Thirteen studies (54%) reported a preparatory information gathering process, where literature reviews (42%, n=10), conversations with community agencies (13%, n=3), or a combination thereof (13%, n=3) provided background to direct creation of the KMb product.

#### Stakeholder Engagement

Most adaptations involved stakeholders through interviews (n=10, 42%) and focus groups (n=7, 29%). A few of the studies engaged groups of participants through interactive workshops^24, 25^ and structured group brainstorming sessions. ^26^ A study that created a prenatal education booklet for Indigenous women in Wyoming initiated the process by building trusting relationships with women from reservations and those attending health conferences. ^27^ These women were then asked to join a volunteer committee to develop culturally appropriate materials. The investigators noted that it took time to build trust with the women individually as well as within the committee.

#### Resources Utilized

Most studies (n=22, 92%) reported some or all of the human resources they utilized during their cultural adaptation process. The involvement of health care professionals was the most prevalent (n=11, 46%), followed by translators (n=8, 33%) and graphic artists or animators (n=7, 29%). Two studies used specialists to moderate their process, one study^24^ had a participatory design expert to assist with guiding exercises and another study^28^ used an experienced moderator to lead their sessions.

#### Evaluation

Nineteen studies (79%) described some form of evaluation for their newly created KMb product, including piloting through surveys, interviews, and focus groups; of these, only 12 (63%) studies reported the results of their evaluation. All studies reporting results focused on evaluating the created or adapted product for use with end-users. Assessments included usability testing, use or knowledge gained through use of the KMb product. However, none of the adaptation processes involved an assessment of end-user engagement and involvement when adapting or creating the KMb product.

### Cultural Adaptations

Details regarding the type of cultural components used and level of stakeholder engagement are presented in Table 3.

**Table 3:**
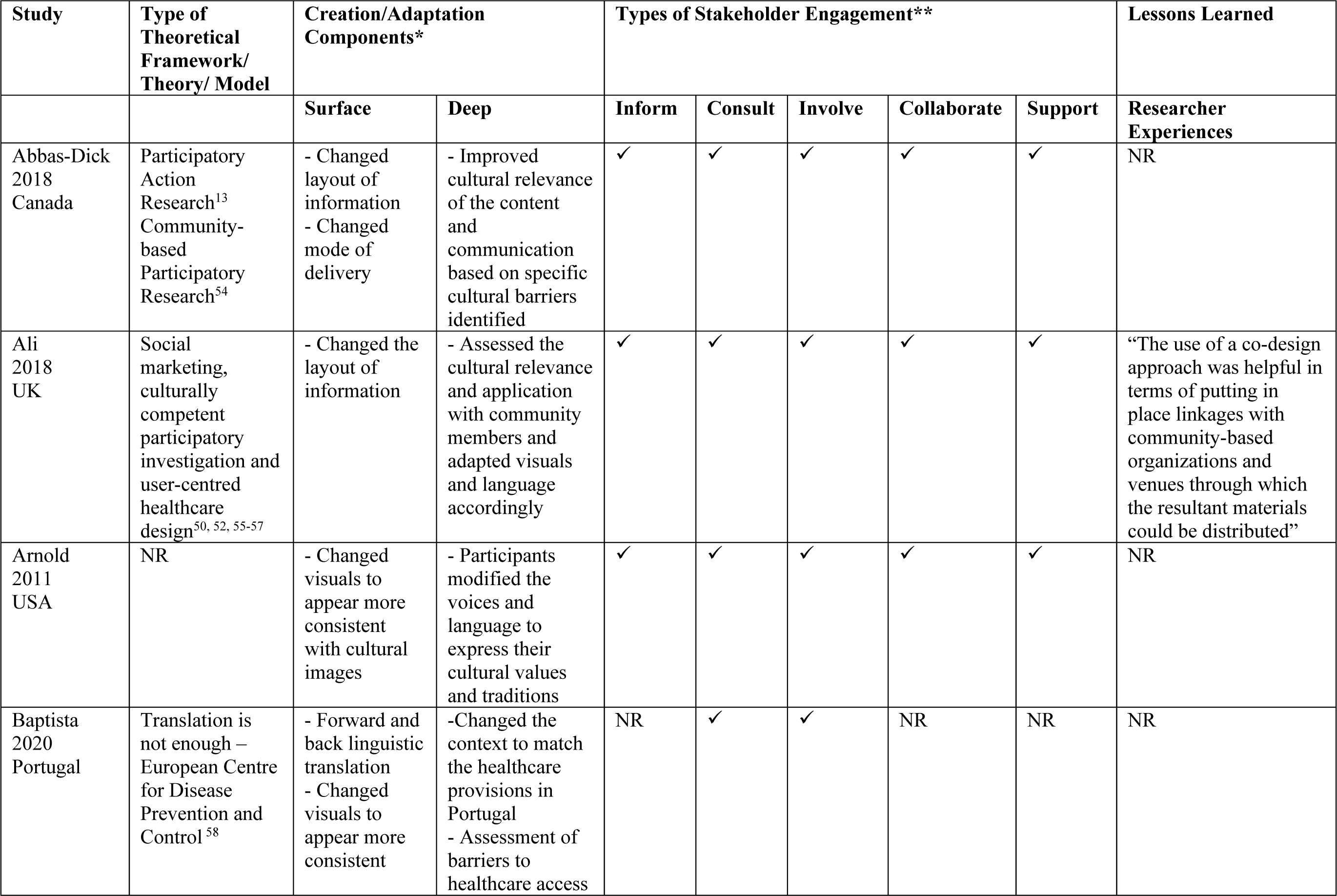
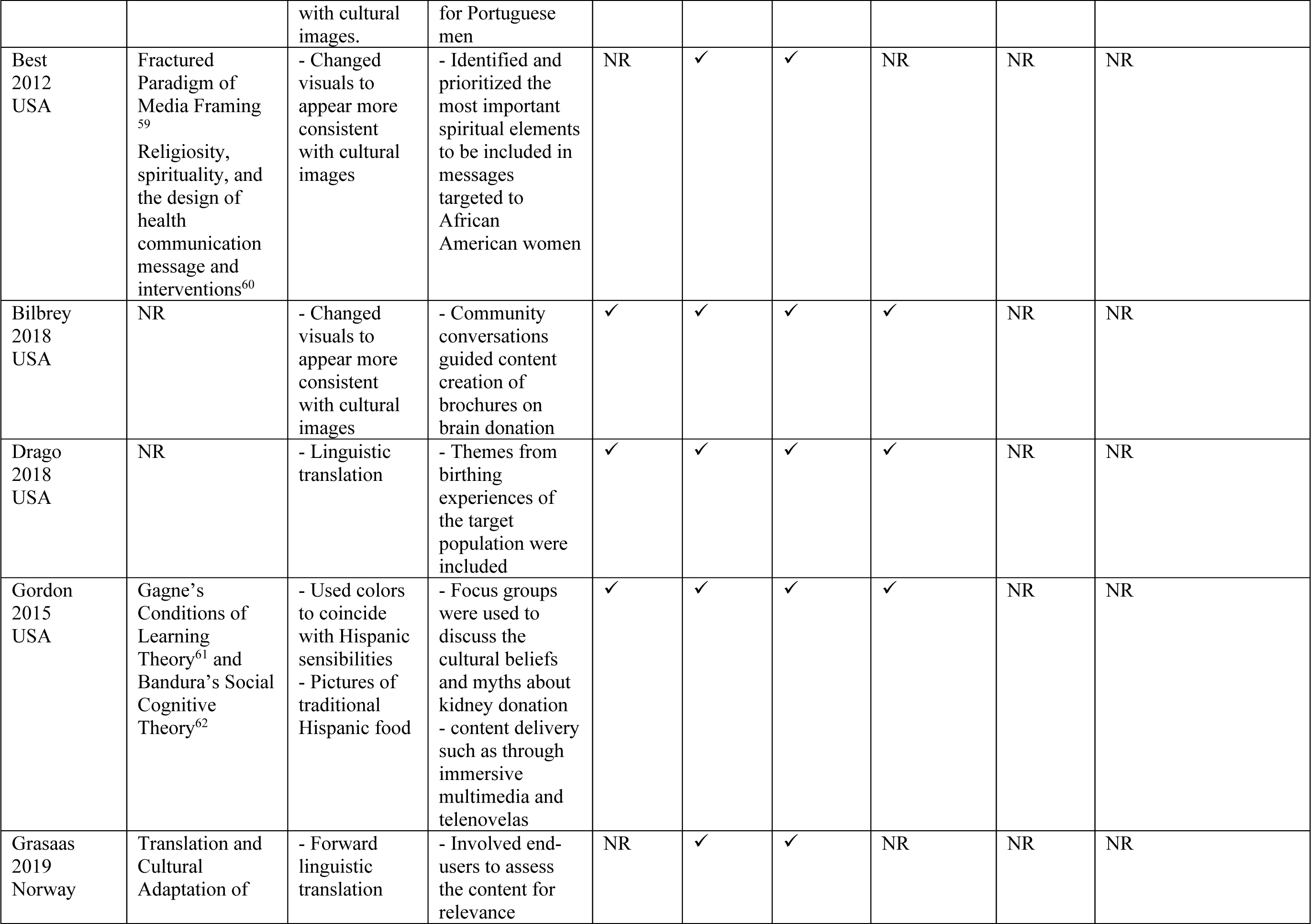
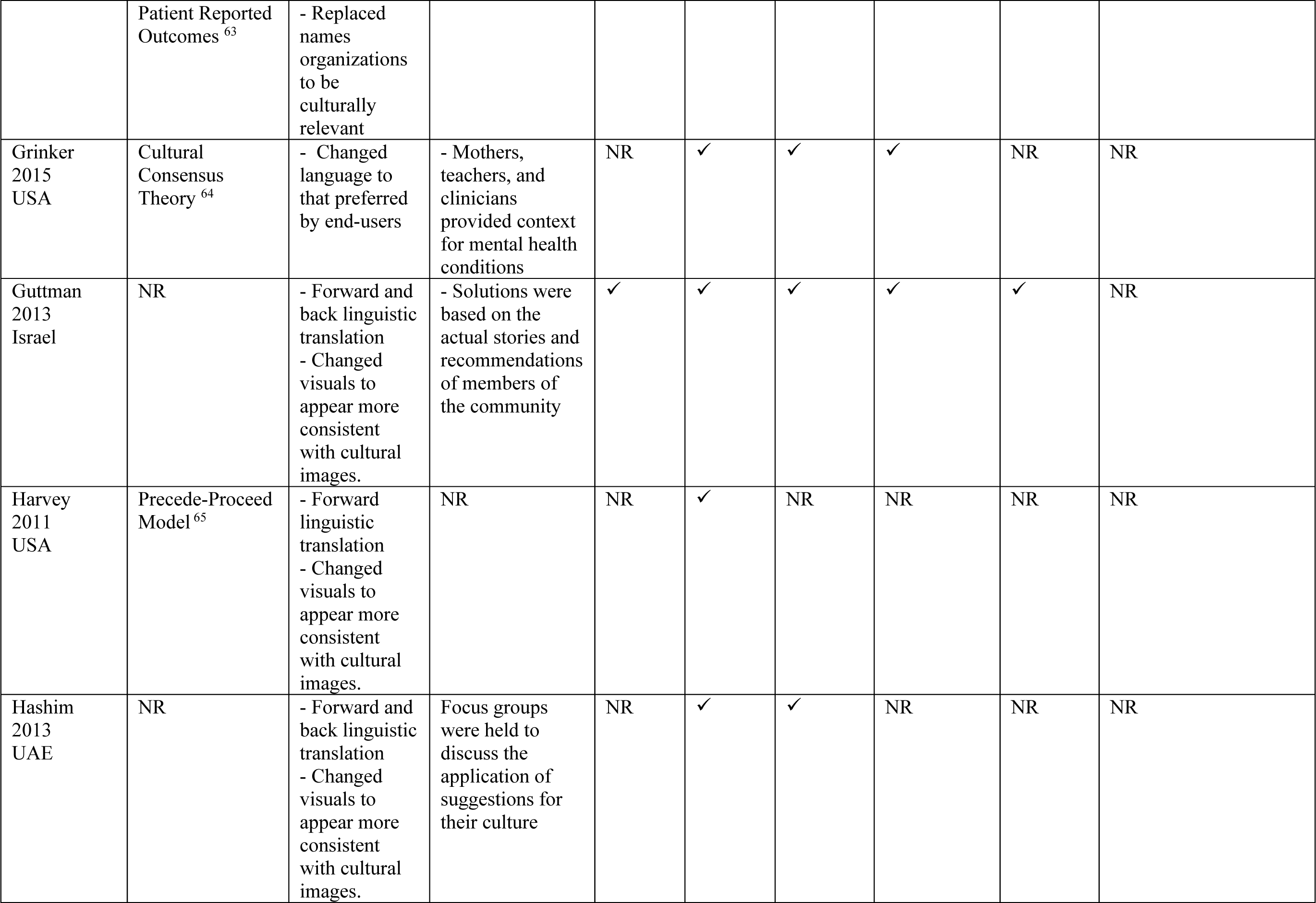
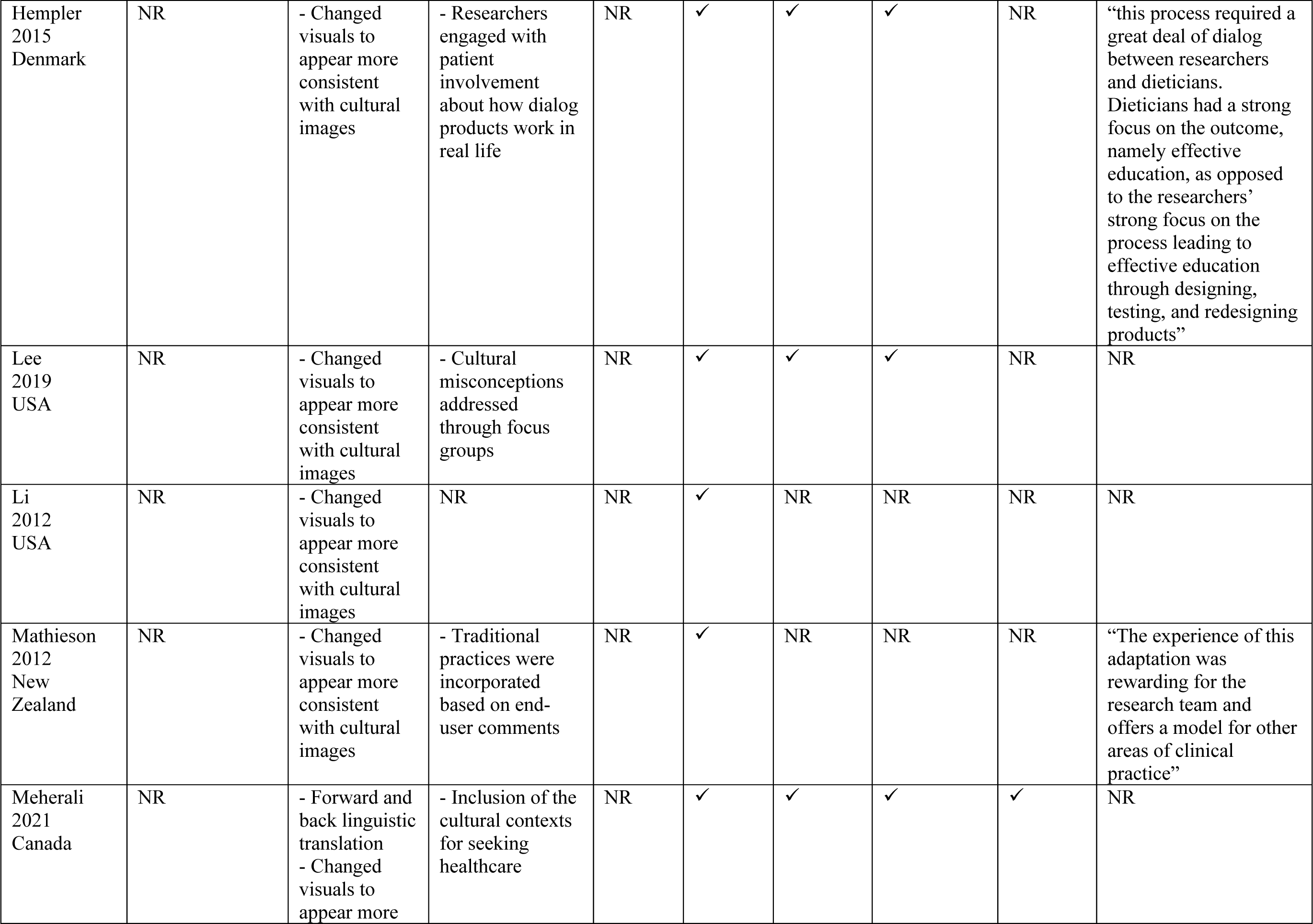
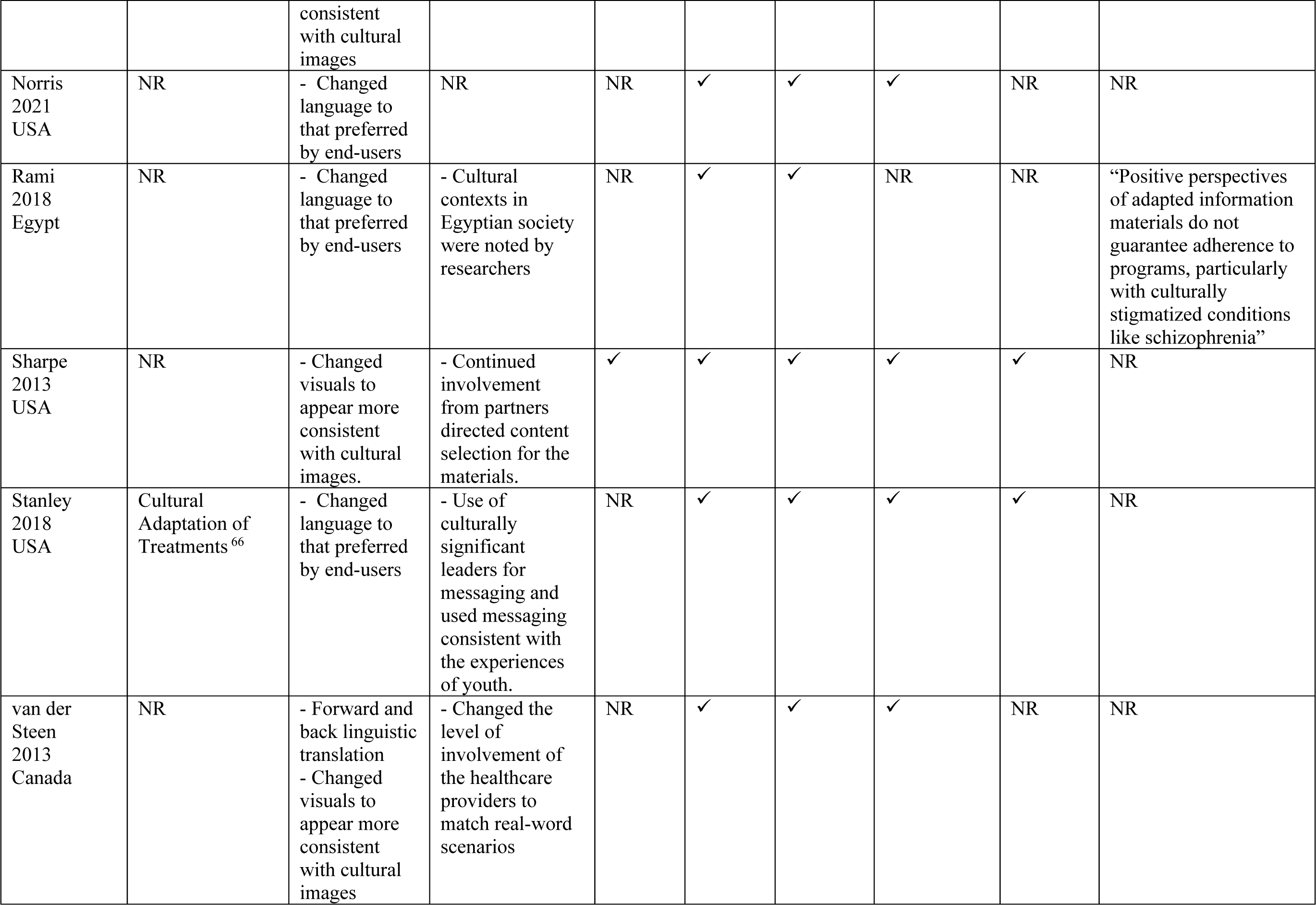
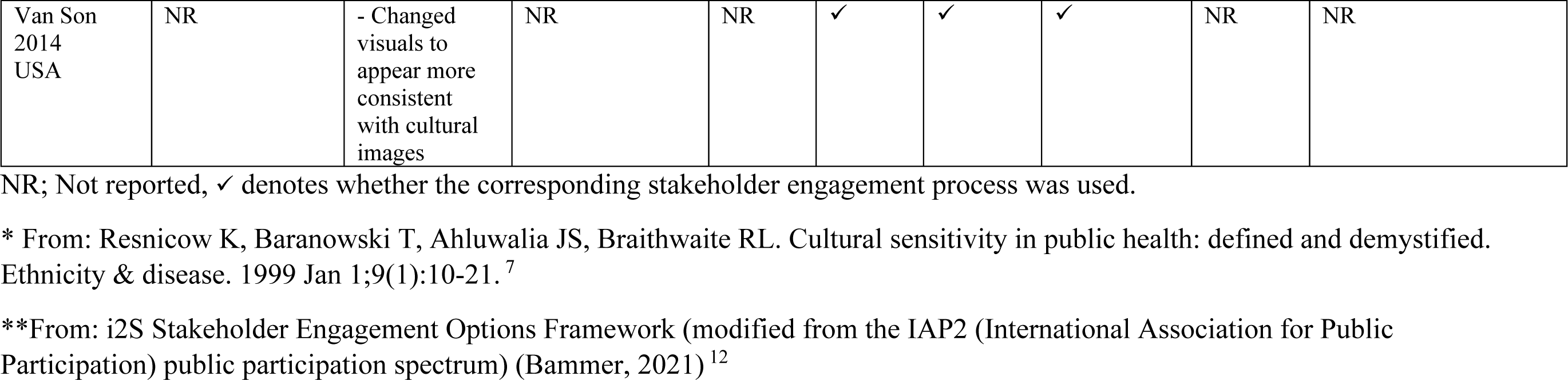
Creation/Adaptation components and stakeholder engagement utilized in included studies.

#### Approach and Type of Adaptation

Two studies (8%) cited using a CBPR^14^ approach, and eight studies (33%) reported using a framework, theory or model to guide their creation or adaptation efforts. All studies utilized surface structure cultural components and most (83%, n=20) included deep structural components in their creation or adaptation processes. To achieve deep structure contextualization authors most commonly consulted specific end-user populations (e.g. clinical populations [21%, n=5], ^25, 29–32^ community members [17%, n=4] ^24, 27, 33, 34^ through focus groups (13%, n=3), ^28, 35, 36^ and assessed various cultural contexts (33%, n=7). ^26, 37–41^

#### Level of Stakeholder Engagement

Five studies^24, 27, 34, 37, 41^ (21%) reported stakeholder engagement approaches representative of all five levels of the i2S version of the IAP2 framework (inform, consult, involve, collaborate, and support). These five studies (21%) created stakeholder committees that were actively involved throughout all phases of the creation or adaptation process. For the remaining 19 (79%) studies, regardless of other stakeholder engagement performed, some form of consultation with stakeholders to learn about cultural considerations were conducted. These consultations were reported to direct deep structure cultural sensitivity considerations for twenty-one of the studies included (88%). With the exception of three^32, 36, 42^ studies (13%), the remaining involved stakeholders throughout the research process, and ten^25, 29, 31, 33, 36, 39, 43–46^ (42%) of the project researchers collaborated with stakeholders. Only two^29, 33^ studies (8%) informed their stakeholders about the research process, and another two^39, 44^ (8%) provided support to stakeholders for implementation and dissemination of the KMb product.

#### Researcher Reflections

Only four (17%) studies included reflections from the research team on the processes for adapting or developing KMb products. Notably, researchers emphasized the importance of forming stakeholder relationships before and throughout the research process, ^24^ that communication between multiple stakeholder groups can be time-consuming, ^25^ that initial positive reception to adapted products does not guarantee adherence to behavior change, ^40^ but that the process of adapting KMb products can be rewarding for researchers (e.g. meeting end-users needs). ^32^

## Discussion

This ScR provides an outline of documented processes used to develop or adapt KMb products for CALD communities, highlights gaps in that literature, and provides direction for future research. As a means of addressing the needs of populations often underserved by health systems, researchers and organizations have begun adapting their KMb products for CALD communities. To the best of our knowledge, this is the first review to synthesize and examine literature on processes and key considerations for adapting or developing KMb products. There appears to be a range of methods employed to address KMb creation for CALD groups. These methods range from original co-created KMb products with participatory frameworks (e.g. Wild et al., 2021 ^5^; Telenta et al., 2020 ^10^) to cultural adaptations of pre-existing KMb products. ^39^ Despite efforts to systematically develop or adapt KMb products, ^39, 47–49^ there is a wide range of cultural and linguistic adaptation processes.

Through this scoping review, we identified 24 studies that reported a variety of methods in adapting and developing KMb products for CALD communities. Across the various cultural communities, modes of information delivery, and approaches/processes cited, only five studies demonstrated deep structure cultural adaptation^50^ and involved end-users in the highest level of stakeholder engagement (as per i2S model). ^12^

Along with study characteristics and creation processes, we extracted information about the depth of creation or adaptation based on Resnicow’s^7^ explanation of surface and deep structures of cultural sensitivity. It has been reported that gaining deep structure cultural knowledge can be a time-consuming process, largely inaccessible to outsiders to the cultural community. ^51^ Although it was not possible to extract information about the cultural background of included studies team members, it is likely that researchers may not identify with the end-user population of study. Researchers who are outsiders to the end-user community lack the necessary information for deep structure cultural sensitivity on their own. However, engaging with community members directly can provide insider perspective for culturally sensitive practice.

Each phase of the i2S framework represents increasing involvement of end-users in research process; ^12^ with the ultimate stage of *Support* representing research decisions led by end-users. The relatively few studies that utilized all five levels of the i2S framework in this review potentially indicate the challenges and commitment required for this process. Studies that exemplified the *Support* phase of the i2S utilized end-user committees that were involved from early conversations about research priorities to eventual dissemination of findings. Ongoing stakeholder engagement is essential to gain insights for deep structure cultural aspects and relevant KMb. ^21^ The majority of studies included in this review engaged in some form of deep structure cultural adaptation, likely due in part to some form of stakeholder engagement consultation^12^ reported in all included studies.

This emphasis on inductive knowledge obtainment and delivery mirrors processes outlined in CBPR^52^ and PAR^13^ frameworks. In both CBPR and PAR, the end-user from the community of study is positioned as a collaborator: someone who has autonomy in the research process as well as insider information for the community of study. ^13, 14, 52^ CBPR has been used as a guiding framework in health intervention literature, and may provide similar guidance for KMb product creation and adaptation. ^53^ Additionally, frameworks used for adapting health interventions, such as the Ecological Validity Model, ^16^ may also offer a systematic approach to cultural adaptations of KMb products. Regardless of the framework used, researchers who choose to develop or adapt KMb products for CALD communities may be well-supported by seeking deep structure cultural understandings through supportive, inductive stakeholder engagement.

A clear gap in the literature was around researchers’ reflections of the processes used, as well as the method of KMb product evaluation. While many studies reported they evaluated the created or adapted KMb product for usability, only one study mentioned evaluating uptake and none assessed impact. Few studies reported the evaluation process or results, and none evaluated the engagement process with stakeholders. Additionally, many did not report on the practicality or feasibility of the processes used (time, resources, engagement), nor whether the product met end-users needed and expectations. Future research should aim to understand the practicalities and nuances of engaging end-users and evaluating the processes to support others in this field. Furthermore, greater transparency by researchers in their adaptation processes would aid in solidifying best practice considerations for culturally adapting KMb products. Ultimately, the most successful methods used by researchers to adapt or develop KMb products for CALD communities could be collated and used to form a framework for future works. This framework could then be evaluated with end-users from various cultural communities to assess its usefulness in this field. However, given how nuanced and tailored KMb should be in meeting the needs of the end-user, perhaps careful planning considering meaningful engagement and being intentional about the best methods to use is key.

### Limitations

This review only included publications in English, yet other cultural creation or adaptation methods studies may be present in languages other than English. The process of defining a KMb product was iterative and largely guided by consensus discussion. The overlap between KMb products and intervention materials was difficult to navigate, particularly when studies did not thoroughly describe their intervention materials.

### Consultation

By examining the methods others have used for their creation and adaptation work, a better understanding around the key considerations when developing or adapting KMb products for culturally and linguistically diverse communities can be achieved.

A methods working group (MWG) stemming from this work will be developed to drive the development of best practices for how to linguistically and culturally develop or adapt KMb products. The MWG will be made up of researchers, cultural knowledge brokers and community members who have firsthand experience and knowledge around how to engage with diverse communities as well as co-design KMb products. By critically evaluating current creation and adaptation practices, we intend to establish a core set of methods and considerations for creating or adapting healthcare decision-making tools for culturally and linguistically diverse communities.

## Conclusion

This review provides information on the various processes and levels of end-user engagement used to culturally develop or adapt a KMb product. While methods and processes, as well as theory or frameworks underpinning the work, varied across projects, it is clear that an important amount of time and resources is required. Significant gaps in the literature still remain surrounding how best to develop or adapt KMb products, what level of engagement is needed, as well as understanding the practicalities of culturally adapting KMb products. Until an appropriate framework exists, researchers would be well-supported by emphasizing cultural sensitivity and meaningful stakeholder engagement in their approaches.

The findings of this review and examples of cultural adaptation could be used to support the development of best practice guidelines for researchers working in this field. Understanding and developing considerations for best practices in this field will assist researchers and organizations in effectively reaching a wider population with health promotion and KMb initiatives.

## Data Availability

Data is available upon reasonable request to the corresponding Author.

## Acknowledgements

We would like to thank Erica Wright for expertise in developing the ScR search strategy. We would also like to thank Sharon Parappally-Joseph, Samantha Cyrkot and Alex Melton for their assistance with screening and data extraction.

## Author Comments

A search update was run in July 2023. Title and Abstract screening is complete, and full text screening and data extraction are in progress. The updated manuscript will be submitted for peer review in a relevant Open Access Journal.

## Supplementary Material A: Eligibility criteria

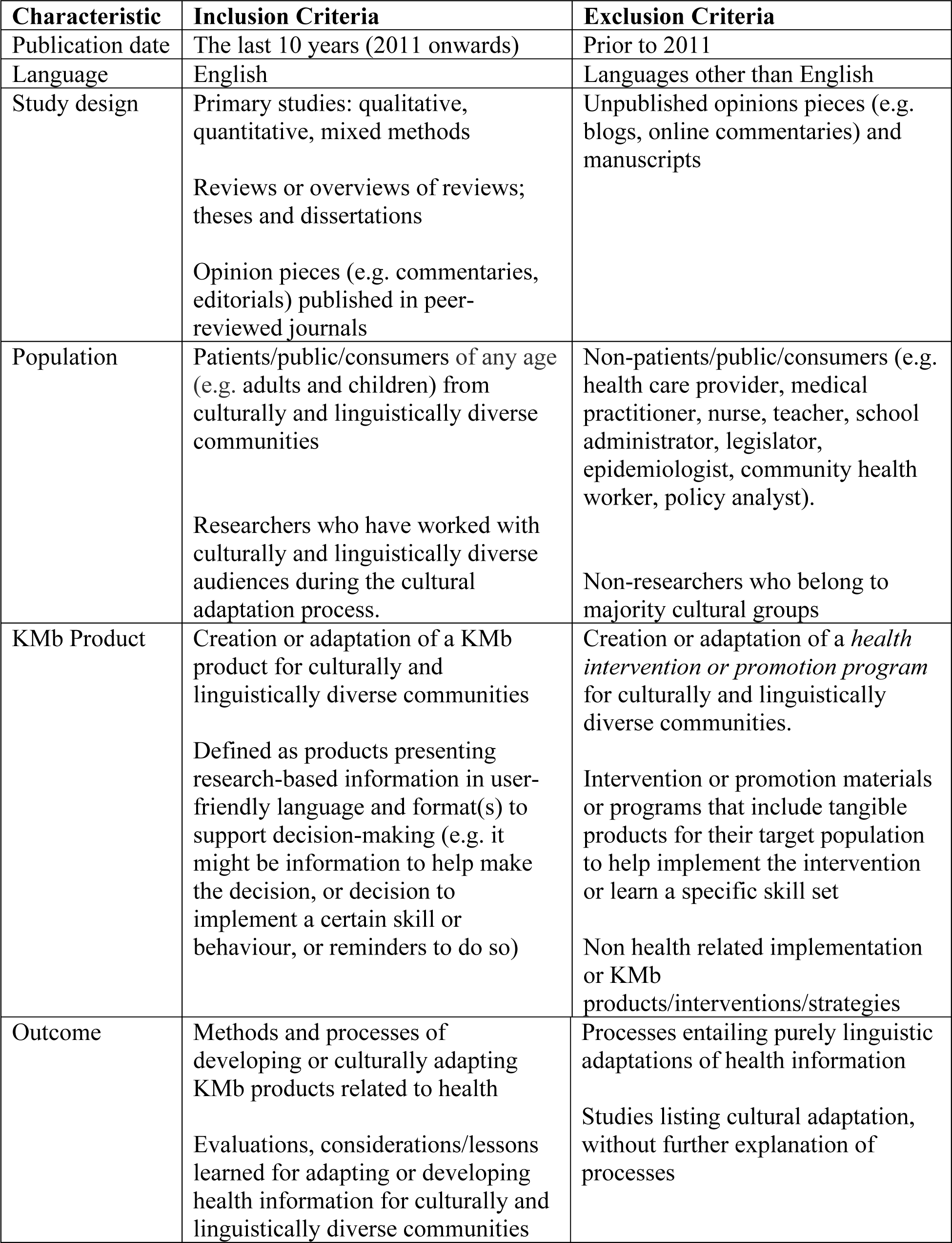
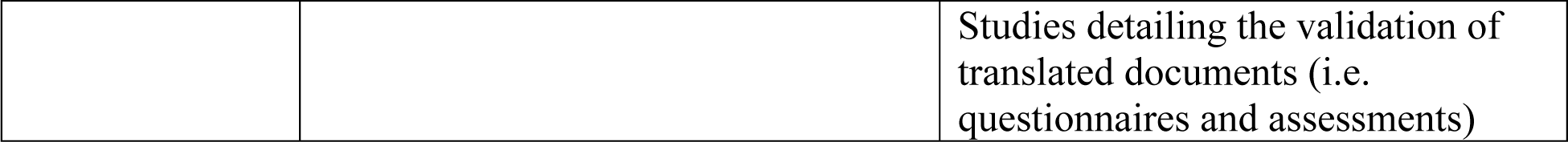

## Supplementary Material B: Medline search strategy

1. Consumer Health Information/ (4095)
2. ((consumer* or patient* or public) adj2 health information).ti,ab,kf. (2194)
3. Health Promotion/ (77181)
4. (health adj2 (promotion* or campaign*)).ti,ab,kf. (40630)
5. Health Communication/ (2815)
6. health communication.ti,ab,kf. (3921)
7. exp Patient Education as Topic/ (87448)
8. ((patient* or consumer*) adj2 education*).ti,ab,kf. (28891)
9. Translational Medical Research/ (11900)
10. ((knowledge adj3 (translat* or transfer* or mobiliz* or mobilis* or exchange or implement* or disseminat* or uptake or adopt* or application* or apply or applie*)) and (service* or tool* or campaign* or program* or resource* or product*)).ti,ab,kf. (10353)
11. ((evidence adj3 (translat* or transfer* or mobiliz* or mobilis* or exchange or implement* or disseminat* or uptake or adopt* or application* or apply or applie*)) and (service* or tool* or campaign* or program* or resource* or product*)).ti,ab,kf. (9326)
12. ((research adj3 (translat* or transfer* or mobiliz* or mobilis* or exchange or implement* or disseminat* or uptake or adopt* or application* or apply or applie*)) and (service* or tool* or campaign* or program* or resource* or product*)).ti,ab,kf. (23587)
13. ((innovat* adj3 (translat* or transfer* or mobiliz* or mobilis* or exchange or implement* or disseminat* or uptake or adopt* or application* or apply or applie*)) and (service* or tool* or campaign* or program* or resource* or product*)).ti,ab,kf. (3574)
14. ((best practice* adj3 (translat* or transfer* or mobiliz* or mobilis* or exchange or implement* or disseminat* or uptake or adopt* or application* or apply or applie*)) and (service* or tool* or campaign* or program* or resource* or product*)).ti,ab,kf. (1510)
15. (“knowledge into practice” or “knowledge to practice” or “knowledge into action” or “knowledge to action”).ti,ab,kf. (5403)
16. (“evidence into practice” or “evidence to practice” or “evidence into action” or “evidence to action”).ti,ab,kf. (13608)
17. (“research into practice” or “research to practice” or “research into action” or “research to action”).ti,ab,kf. (12970)
18. (“innovation* into practice” or “innovation* to practice” or “innovation into action” or “innovation to action”).ti,ab,kf. (251)
19. (evidence based adj2 (program* or service* or intervention* or campaign* or tool* or resource*)).ti,ab,kf. (11038)
20. (KMb adj2 (product* or tool* or intervention* or strateg* or campaign* or service*)).ti,ab,kf. (306)
21. (brochure* or booklet* or pamphlet*).ti,ab,kf,hw. (10594)
22. ((app or apps or application*) adj3 (mobile or device* or cell* or iphone* or android* or smart-phone* or smartphone*)).ti,ab,kf,hw. (46688)
23. (social media or facebook or instagram or twitter).ti,ab,kf,hw. (23959)
24. or/1-23 (367959)
25. Culture/ (33657)
26. cross-cultural comparison/ (26456)
27. Cultural Characteristics/ (16714)
28. Cultural Competency/ (5981)
29. Culturally Competent Care/ (1823)
30. cultural diversity/ (12046)
31. ((cultur* or ethnocultur*) adj5 (adapt* or translat* or modif* or tailor*)).ti,ab,kf. (21204)
32. ((transcultur* or crosscultur* or multicultur*) adj5 (adapt* or translat* or modif* or tailor*)).ti,ab,kf. (533)
33. ((cultur* or ethnocultur* or transcultur* or crosscultur* or multicultur*) adj5 (divers* or competen* or appropriat* or responsive or relevan* or characteristic*)).ti,ab,kf. (38389)
34. ((cultur* or ethnocultur*) adj4 (specific* or sensitiv* or inclusiv*)).ti,ab,kf. (29812)
35. ((ethnic* or minority or minorities) adj5 (adapt* or translat* or modif* or tailor*)).ti,ab,kf. (1526)
36. ((ethnic or minority or minorities) adj2 (communit* or group*)).ti,ab,kf. (45850)
37. (CALD adj2 (communit* or group* or background*)).ti,ab,kf. (161)
38. ((cultur* or ethnocultur* or transcultur* or crosscultur* or multicultur*) adj2 linguist*).ti,ab,kf. (2995)
39. ((co-design* or co-creat* or codesign* or cocreat*) and (cultur* or ethnic* or ethnocultur* or transcultur* or crosscultur* or multicultur*)).ti,ab,kf,hw. (407)
40. ((participatory adj2 (design* or research or framework* or method*)) and (cultur* or ethnic* or ethnocultur* or transcultur* or crosscultur* or multicultur*)).ti,ab,kf,hw. (2672)
41. (action research and (cultur* or ethnic* or ethnocultur* or transcultur* or crosscultur* or multicultur*)).ti,ab,kf,hw. (755)
42. or/25-41 (200866)
43. 24 and 42 (11303)
44. (patient* or consumer* or audience* or end user* or service user* or client* or public or parent* or mother* or father* or caregiver* or care giver*).ti,ab,kf,hw. (8730104)
45. 43 and 44 (6725)
46. limit 45 to english language (6510)

